# Viral infectivity in pediatric SARS-CoV-2 clinical samples does not vary by age

**DOI:** 10.1101/2022.10.17.22281193

**Authors:** Madaline M. Schmidt, Hannah W. Despres, David J. Shirley, Michael E. Bose, Kate C. McCaul, Jessica W. Crothers, Kelly J. Henrickson, Benjamin Lee, Emily A. Bruce

## Abstract

During the early months of the SARS-CoV-2 pandemic, notable uncertainty emerged regarding the role of children in transmission dynamics ^1^. With time, it became more clear that children were susceptible to infection with SARS-CoV-2, but that the vast majority of children experienced mild symptoms with lower incidence of severe disease ^2^. This pattern remained consistent despite the later emergence of SARS-CoV-2 variants, including Delta and Omicron, even among children <5 ineligible for vaccination^3^. The relative lack of severe disease in the pediatric population raised questions regarding viral kinetics and infectivity in children versus adults.

## INTRODUCTION

We hypothesized that unique virologic features in children could explain this apparent decrease in symptoms and transmissibility early in the pandemic. Due to the challenges posed by measurement of infectious viral titers, the majority of work examining viral loads in clinical samples has measured viral RNA levels, as determined by RT-qPCR cycle threshold [C_T_]. A previous study using this technique reported no differences in viral RNA load in adults and children, when controlling for the presence of symptoms ^4^. A different study reported both RNA viral load and level of infectious virus using a semi-quantitative method (TCID_50_) in pediatric clinical samples ^5^. In contrast however, other work indicates that ancestral SARS-CoV-2 replicates less efficiently in both children and pediatric versus adult nasal epithelial cells, a defect that Omicron was able to abolish ^5-7^. Finally, we and others have demonstrated a dynamic relationship between C_T_ values and infectious viral titers with potential for significant discrepancies and a ratio dependent on both viral and host factors, ^8^ but this work did not include children ^9^.

Therefore, to further understand SARS-CoV-2 infection in children, we investigated the ratio of infectious virus titer to RNA viral load in children aged 0 to <18 years old. We hypothesized that the ratio of infectious virus to RNA viral load would be positively associated with age.

## METHODS

### Sample Selection

Banked SARS-CoV-2 positive nasopharyngeal specimens from children 0 to <18 years old collected and stored at Children’s Wisconsin, Milwaukee, Wisconsin between September 14, 2020 and May 17, 2021 were identified. Deidentified samples were binned into four age groups (<1, 1-5, 6-11, and 12-17) and stratified by clinical C_T_ value (<20, 20-24, 25-29, and 30-34) to select a sample of children representing the full spectrum of both age and C_T_ value. The study received an exempt determination for use of deidentified specimens from the University of Vermont (UVM) Institutional Review Board and the Children’s Wisconsin Institutional Review Board.

### RNA extractions and RT-PCR

Total nucleic acid was extracted on the NucliSENS easyMAG or EMAG automated extraction instruments (bioMerieux). SARS-CoV-2 RNA was detected using previously published primers/probes for the SARS-CoV-2 E gene (Sarbeco^10^) on the 7500 Fast Real-Time PCR System or QuantStudio 7 Pro platforms.

### Viral Titrations

SARS-CoV-2 viral titering was conducted under BSL-3 conditions at UVM using a microfocus forming unit (FFU) assay in VeroE6-TMPRSS2 cells, which increases assay sensitivity compared to standard VeroE6 cells, as previously described ^8^.

### Statistical Analysis

Viral titers were log-transformed for analysis. Linear regression was used to predict log titer as a function of C_T_, fitting separate models without age and to control for continuous and categorical age effects. Models were compared by F test. Data were analyzed and plotted with R. Code is available at https://github.com/emilybrucelab.

## RESULTS

N=144 clinical specimens were selected to determine the relationship between the infectivity of SARS-CoV-2 in pediatric samples and RNA viral load. As expected, higher RNA viral load generally correlated with higher infectious virus titer, although as reported previously this ratio was somewhat variable ^8,9^. In linear regression, the relationship between infectious viral titer and C_T_ was not significantly modified by age (P=0.156) or age group (P=0.355 overall by F test). These data indicate that there is no difference in the infectiousness of SARS-CoV-2 produced by children, regardless of age.

## DISCUSSION

Consistent with previous findings, we found no significant differences in the relationship between SARS-CoV-2 infectious virus titer and RNA viral load in children across the pediatric age spectrum ^4,7^. Our findings suggest equal levels of viral infectivity in children and in adults with similar RNA viral loads. Limitations of this study include lack of access to viral sequencing and individual level metadata, which could reveal differences in infectivity as a result of viral genetic background, days post-symptom onset, host immune status, and vaccination status. Furthermore, there was no direct comparison with adult samples, although we did include samples in older teens who would closely resemble adults biologically.

## Data Availability

Code is available at https://github.com/emilybrucelab. All data produced in the present study are available upon reasonable request to the authors.

## Abbreviations

SARS-CoV-2: (severe acute respiratory syndrome coronavirus 2)
COVID-19: (coronavirus disease 2019)
C_T_: (cycle threshold)
RT-qPCR: (reverse transcription-quantitative polymerase chain reaction)
RNA: (ribonucleic acid)
TMPRSS2: (transmembrane protease, serine 2)
FFU: (focus forming unit)

## ACKNOWLEDGEMENTS

We thank Ms. Kubinski and Dr. Oetjen for technical assistance.

---

**Figure 1.**
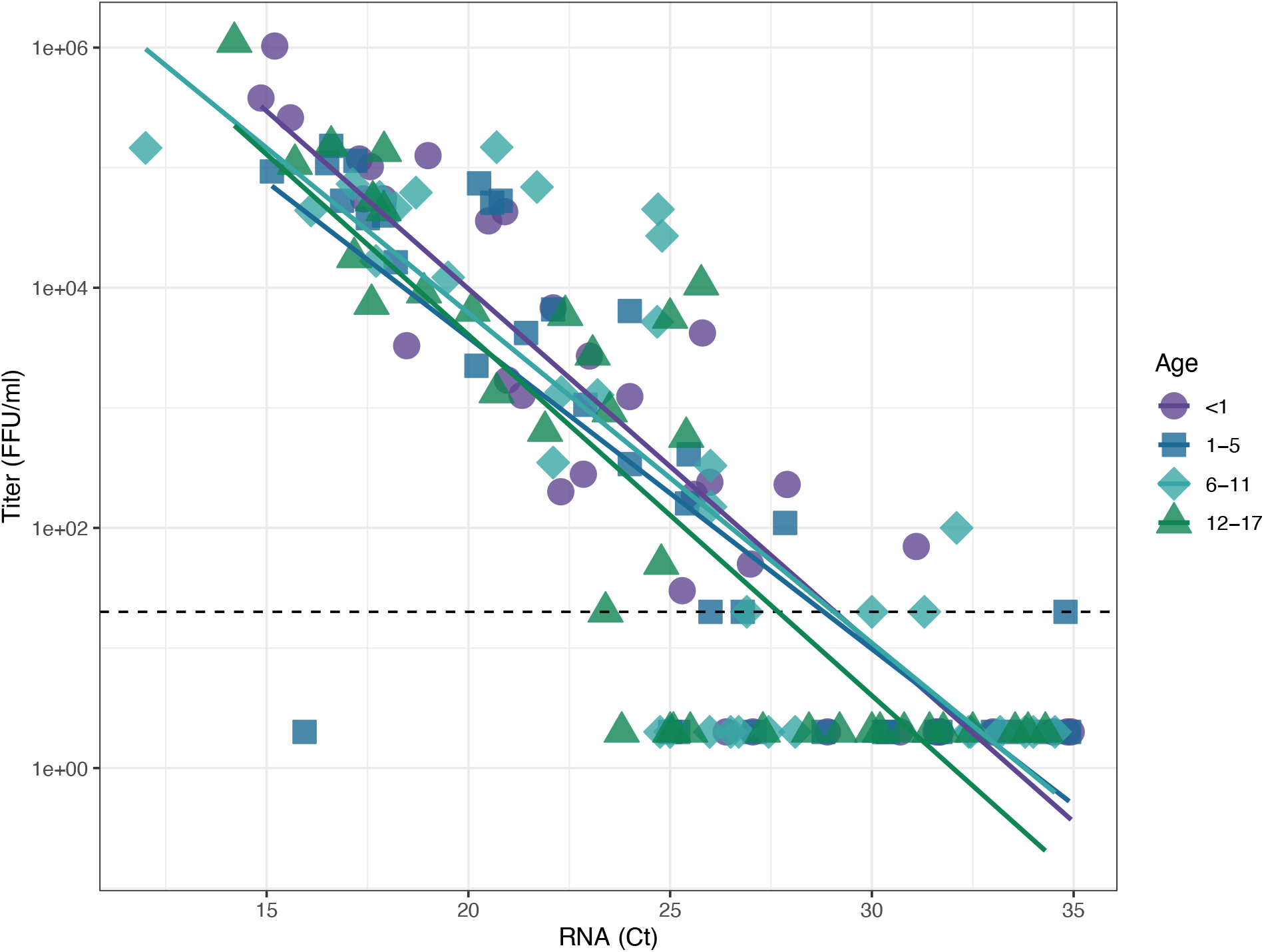
SARS-CoV-2 viral infectivity does not vary by age in a pediatric population. A set of 144 clinical samples from children infected with SARS-CoV-2 was used to examine the relationship between infectious virus titer and RNA viral load as a function of patient age. Individual specimen measurements of E gene RNA levels (C_T_) on the x-axis are plotted against viral titer, as measured in focus forming units (FFU/mL) on the y-axis. Dashed line indicates the limit of detection for infectious titer (20 FFU/mL). Samples for which we could not measure a viral titer were assigned fixed values of one-tenth the limit of detection (2 FFU/mL). Lines of best fit were generated by linear regression on log-transformed titer data as a function of C_T_ and age group.

